# Using Independent Components Analysis To Identify Breast Cancer Using Dynamic Active Thermography

**DOI:** 10.1101/2025.11.23.25340812

**Authors:** Meir Gershenson, Jonathan Gershenson

## Abstract

Mammography is the gold standard for breast cancer detection, but there remains a need for supplementary techniques to find lesions it misses. This research explores dynamic far-infrared thermography, which uses a thermal camera to monitor changes in skin temperature over time. The study aims to characterize the thermal behavior of the vascular system, both at rest and in response to a temperature stimulus that causes Vaso modulation (blood vessel constriction or dilation). We use matrix factorization to reduce 20 thermal frames and the static frame into a final set of three frames and a time-trace matrix. The results yielded clear images of these thermal and vascular responses. Using the time matrix without classification into sick/healthy, I produce an algorithm with output of three parameters. With apparent clustering correlated with the patient classification as sick or healthy. The authors recommend further studies with more extensive clinical data to validate this new approach.

## INTRODUCTION

Breast cancer is one of the most diagnosed and deadly cancers among women, making early detection essential for improving outcomes [1][2]. The primary modalities for breast cancer screening and diagnosis include mammography, ultrasound, and MRI. While mammography is the gold standard, its sensitivity decreases with increasing breast density, and there are concerns about potential iatrogenic-related malignancies. Moreover, mammography remains cost-prohibitive in developing countries, highlighting the need for complementary imaging methods. Thermography, which analyzes infrared radiation emitted by the body based on temperature, offers a cost-effective, beneficial alternative. It creates an overall map of the skin’s temperature, achieving a thermal sensitivity of 0.02 °C or better. However, the utility of previous thermography systems has been limited by low medical sensitivity and a lack of extensive population studies, prompting the FDA to issue warnings against their prior use for breast cancer screening [3].

Tumors typically exhibit an increased metabolism and blood supply due to angiogenesis, which can lead to temperature differences observable in thermal imaging. Although IR imaging can help identify neovascularization, it has not yet demonstrated a direct correlation between blood vessel heat signatures and tumor locations.

Innovative techniques, such as “dynamic thermal imaging”(DTI) [4], enhance diagnostic accuracy by capturing images under cold-stress conditions. To perform standard DTI, the body is cooled using a stream of air after a period of acclimatization. An infrared camera then records the thermal images. Typically, the temperature is recorded at the end of the cooling process [5]. As we record the temperature at the end of the cooling period, we observe relaxation of vasodilation.

Vasodilation is the physiological process of actively increasing the diameter of blood vessels, primarily arteries and arterioles, to regulate blood flow and pressure within the body. It is a critical physiological process that regulates body temperature, blood pressure, and the distribution of blood flow. Immediate local responses and delayed systemic (remote) hormonal signals control the vascular modulation. In a previous work, “Dynamic Vascular Imaging Using Active Breast Thermography” [6], we introduced an advanced detection method that analyzed changes in blood vessel temperature following controlled cooling. This technique focuses on the active physiological response of vasodilation to differentiate tumor-related heat patterns from surface blood flow. While the method identified vasodilation in the vascular system, it did not detect cancer. Another tentative interpretation method is reverse modeling using a static thermal image to infer heat flow from the tumor [7] [8] [9]. That method was successful. The significant disadvantage is the substantial thermal signal from the vascular system, with approximately 90 percent of the tumor heat carried by the vascular system and only 10 percent by the tissue [10]. This vascular heat appears remotely at the skin and can mask or even overshadow tissue heat. Neovascularization is a key indicator of malignant breast tumors because cancer requires a new, accelerated blood supply to grow and metastasize [11][12][13][14]. While its presence is not exclusive to cancer, a high density of new, abnormal blood vessels is strongly associated with aggressive breast cancer and poor prognosis. With advances in technology, recent studies have also used AI to interpret breast thermograms [15][16][17][18][19][20][21][22][23]. One problem with AI is the “black box” issue: the internal workings of many AI systems are opaque, making it difficult for clinicians to understand how the system arrived at its conclusions. AI’s lack of clarity and verifiability can reduce trust in AI-generated decisions. AI techniques likely identify neovascularization among other undetermined factors.

Matrix factorization [24] converts complex thermal image data, represented as a large matrix ***V***, composed of individual vectors, each representing a single frame, into two smaller, more manageable matrices: a basis matrix ***W*** and a coefficient matrix ***H (V = H*W)***. The basis matrix ***W*** contains the core “thermal basis vectors,” which represent the most significant thermal patterns found across the entire image sequence. The coefficient matrix *H* then serves as a set of weights, indicating how these core patterns combine at each time step to reconstruct the original data. This decomposition effectively separates prominent thermal signatures from background noise, enabling more precise detection of subtle temperature changes. The common factorization used for thermal breast screening is Principal Component Analysis (PCA)[25], Independent Component Analysis, and Non-Negative Matrix Factorization (NNMF)[26][27], with some variations of NNMF. Notable review articles on previous DTI methods for breast cancer detection exist [28][29][30][31]

## II Method

In a previous study using virtual waves, we isolated heat reflection from the vascular system and identified vasodilation in arteries and veins [6]. The study failed to achieve its primary goal of distinguishing tumor heat from that of the nearby vasculature. The virtual wave used in the previous work is transparent to static and quasistatic heat propagation, such as constant metabolic heat and systemic vasodilation. In this work, we used techniques developed in an earlier work, but without the virtual wave. To overcome the small number of well-characterized patients, we included all patients who didn’t fail due to excess motion. We trade ‘larger statistic’ for ‘quality,’ which was justified because it led to an algorithm that could identify possible cancer.

### A. Goal

In a previous study using virtual waves, we isolated heat reflection from the vascular system and identified vasodilation in arteries and veins [6]. The study failed to achieve its primary goal of distinguishing tumor heat from that of the nearby vasculature. The virtual wave used in the previous work is transparent to static and quasistatic heat propagation, such as constant metabolic heat and systemic vasodilation. In this work, we used techniques developed in an earlier work, but without the virtual wave. To overcome the small number of well-characterized patients, we included all patients who didn’t fail due to excess motion. We trade ‘larger statistic’ for ‘quality,’ which was justified because it led to an algorithm that could identify possible cancer.

### B Approach

In a previous study, an analysis of skin DTI data after external cooling isolated the heat signature of veins, which exhibited both passive diffusion-based and active physiological responses [6]. Using techniques borrowed from thermal nondestructive evaluation (NDE), the researchers employed Virtual Wave Transformation (VWT) and matrix factorization methods, such as PCA and ICA, to process the data and distinguish these responses. The limitation of VWT is that it detects and resolves diffusion propagation but remains transparent to slow-varying heat responses, such as delayed systemic responses. To overcome this limitation, I performed matrix factorization, but without VWT. Eliminating the VWT reduces time resolution but increases sensitivity to the delayed responses. We can apply VWT to the diffusive components later. NNMF is a standard matrix factorization method used in DTI analysis [32]. Its use assumes that the DTI signal is positive and contains no negative components. As we have seen in previous work using VWT, individual reflections can be negative while the overall response remains positive. Using ICA, we separate the data into its main components, W, and a time matrix, H. The ICA algorithm is inefficient because it relies on random search. PCA analysis is much more efficient as it has an analytical implementation. We used PCA factorization as an initial approximation for the ICA algorithm. Doing so makes it more efficient and fixes the component ordering, which varies in a random search of the ICA. We use the time matrix H for additional analysis.

Units

## III DATA ANALYSIS

### A Image ICA Analysis

By stacking all columns for each patient, we created two matrices: the first containing only the region of interest (pixels within the breast), and the second containing the whole frame surrounding the breast. We used the second set for image display, and the first set for analysis. We applied the results of the first set to the complete image. For each group of images (one breast), we apply PCA, then ICA. Three components were sufficient. Figures 1 and 2 are of patient T016, who is healthy. Figure 1a is of the raw static image, while Figures 1b to 1d are of the three component images. The first one—the vascular system—is almost identical to the static frame. The other two components are much smaller in magnitude. Figure 2 is the matrix H. The first 20 points correspond to the dynamic data, and the 21st point is of the static frame. In this, the third component is significant only at the static frame; it corresponds to a slow physiological response.

**Figure 1.**
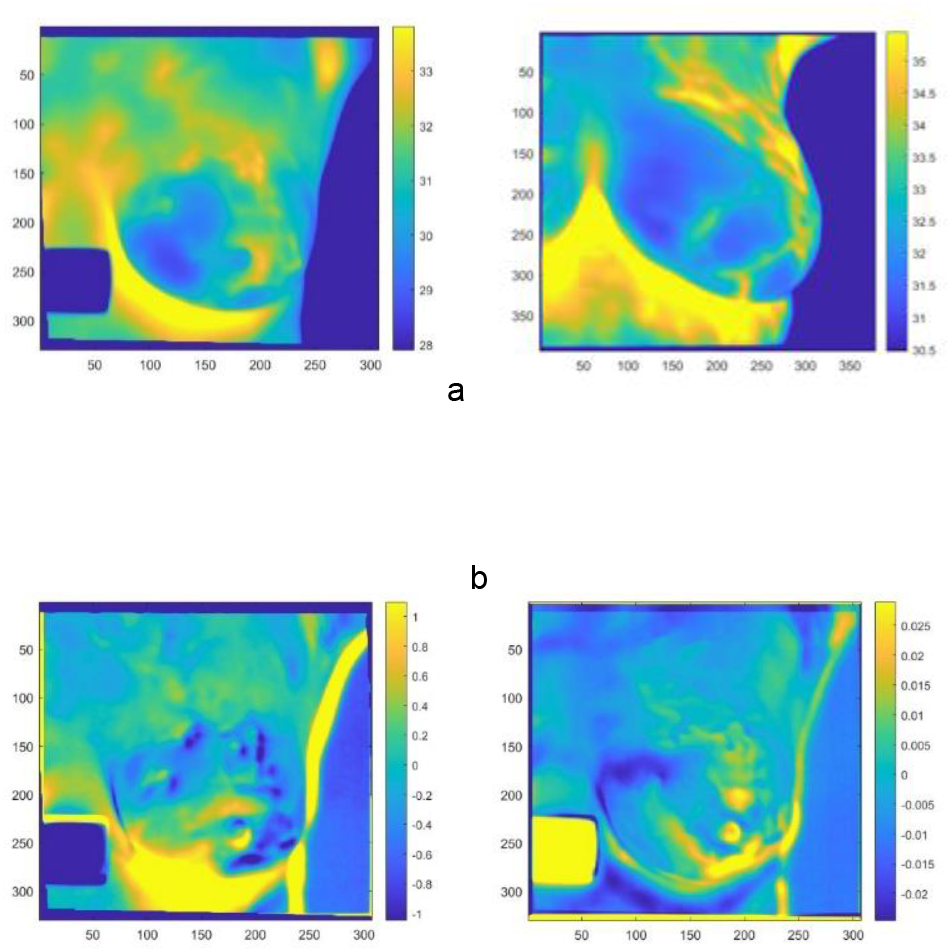

**Figure 2.**
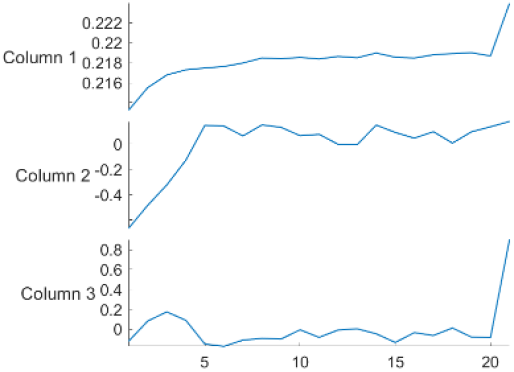
Patient T016. Time matrix H. Each trace corresponds to one image. Points 1-20 are the dynamic amplitude, and 21 is the static amplitude.

Figures 3 and 4 show patient T282, who is ill. Figure 3a shows the image of the raw static image, while Figures 3b to 3d show the three components with the amplitude of the static frame. The first component is of the vascular system and is almost identical to the static frame. We are not sure about the interpretation of the second component. Figure 4, second trace, identifies it as related to vasodilation and as not contributing to the static frame. Figure 3d, which differs from the healthy patient: the slow physiological signal is much larger in amplitude, 1.50 °C for the sick patient versus 0.025 °C for the healthy patient. A hot spot is in the lower inner quadrant, the same as in the cancer diagnosis. There is a shadow across the hot region of Figure 4d, at the same position as a hot line of Figures 3a or 3b. Figures 3, 5, 6, and 7 show the four patients with identified tumor locations.

**Figure 4.**
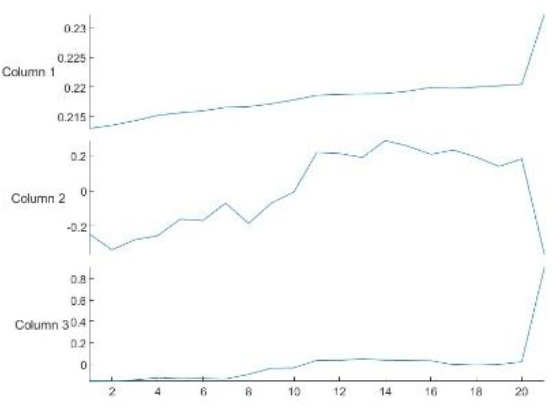
Time matrix of Patient T282*H*. Each trace corresponds to one image. Points 1-20 are the dynamic amplitude, and 21 is the static amplitude.

**Figure 5.**
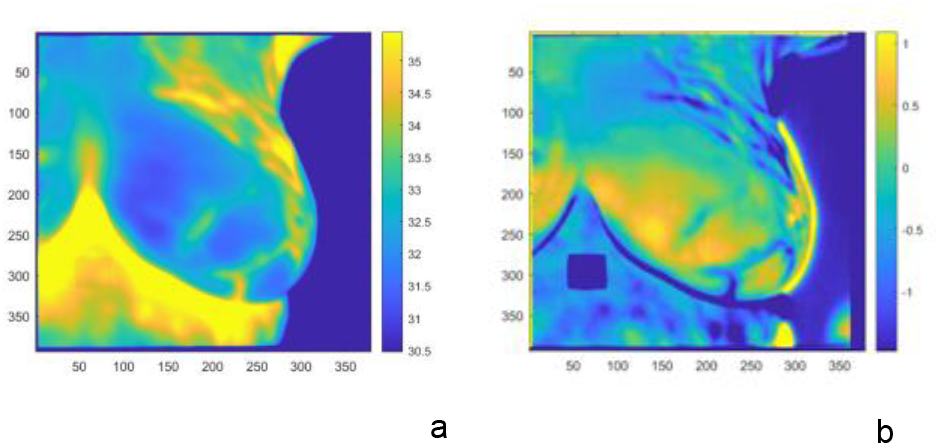

**Figure 6.**
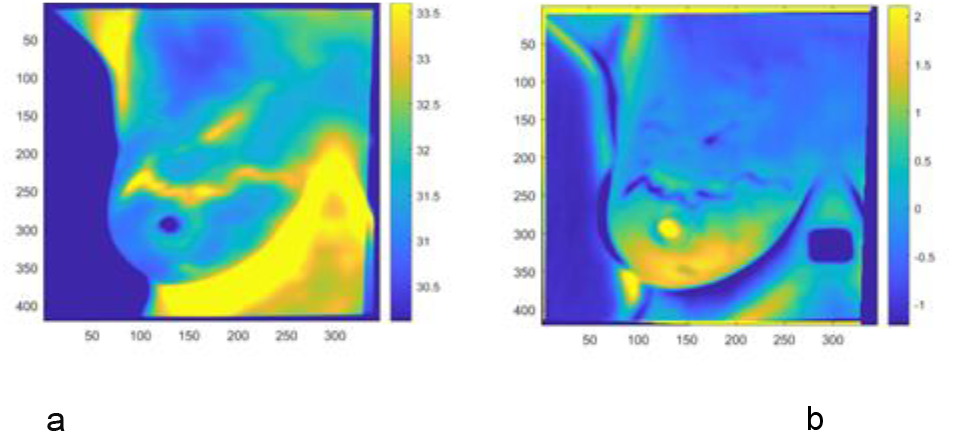

**Figure 7.**
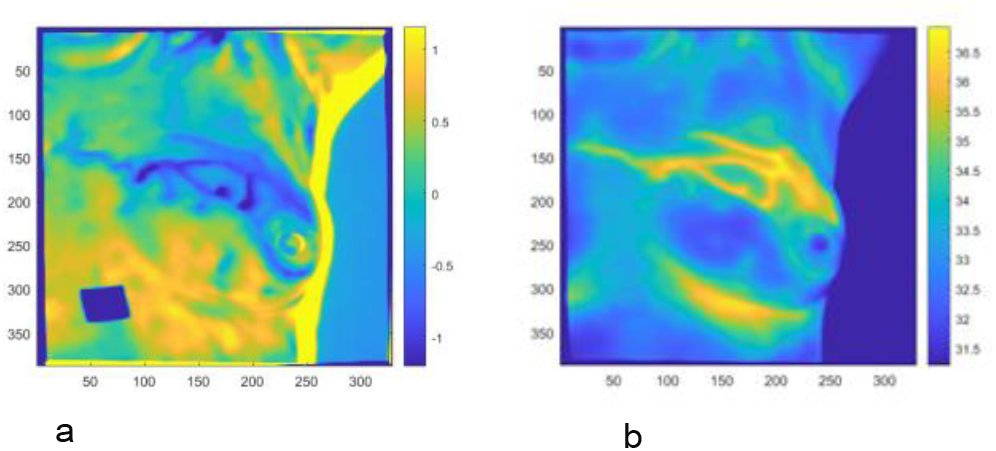

### B Time Signature Analysis

In the previous section, I visually analyzed the three components of the image matrix W to identify suspected internal tumor heat. In this section, I developed tools to analyze the time-matrix H. For this analysis, we used the entire data set without labeling it as sick or healthy; the healthy/sick information was used only at the final verification. As such, it reduces concerns about bias when using the same data for both training and validation. Initially, we tried using both healthy and sick datasets separately, which did not affect the results. The ICA algorithm normalizes each column of the matrix, H, to 1 and carries no temperature information; we reintroduce the temperature information by multiplying it by the norm of the corresponding image.

We reduced the 3×21 matrix ***H*** for each patient to a single vector of 12 parameters by using only the first, 10th, and 20th columns, along with the static frame, point 21. We smoothed each point using the two adjacent points on each side. We stack the 374 × [1×12] vectors into a single [374×12] matrix. The ICA algorithm identified a [12×4] mixing matrix. Four columns were optimal, as the 5th column was almost identical to the previous one. We used this [12×4] matrix for discrimination. A 1×12 vector represents each patient. By multiplying this vector by the discrimination matrix, we generate four numbers. We plot all patients as red dots for healthy patients and black dots for sick patients (this is the first time in the process that we distinguish between the two populations). The first parameter has no diagnostic value.. An additional application of ICA to the reduced three-parameter mixing matrix (excluding the first nondiscriminating row) will realign it as shown in Figure 8.

**Figure 8.**
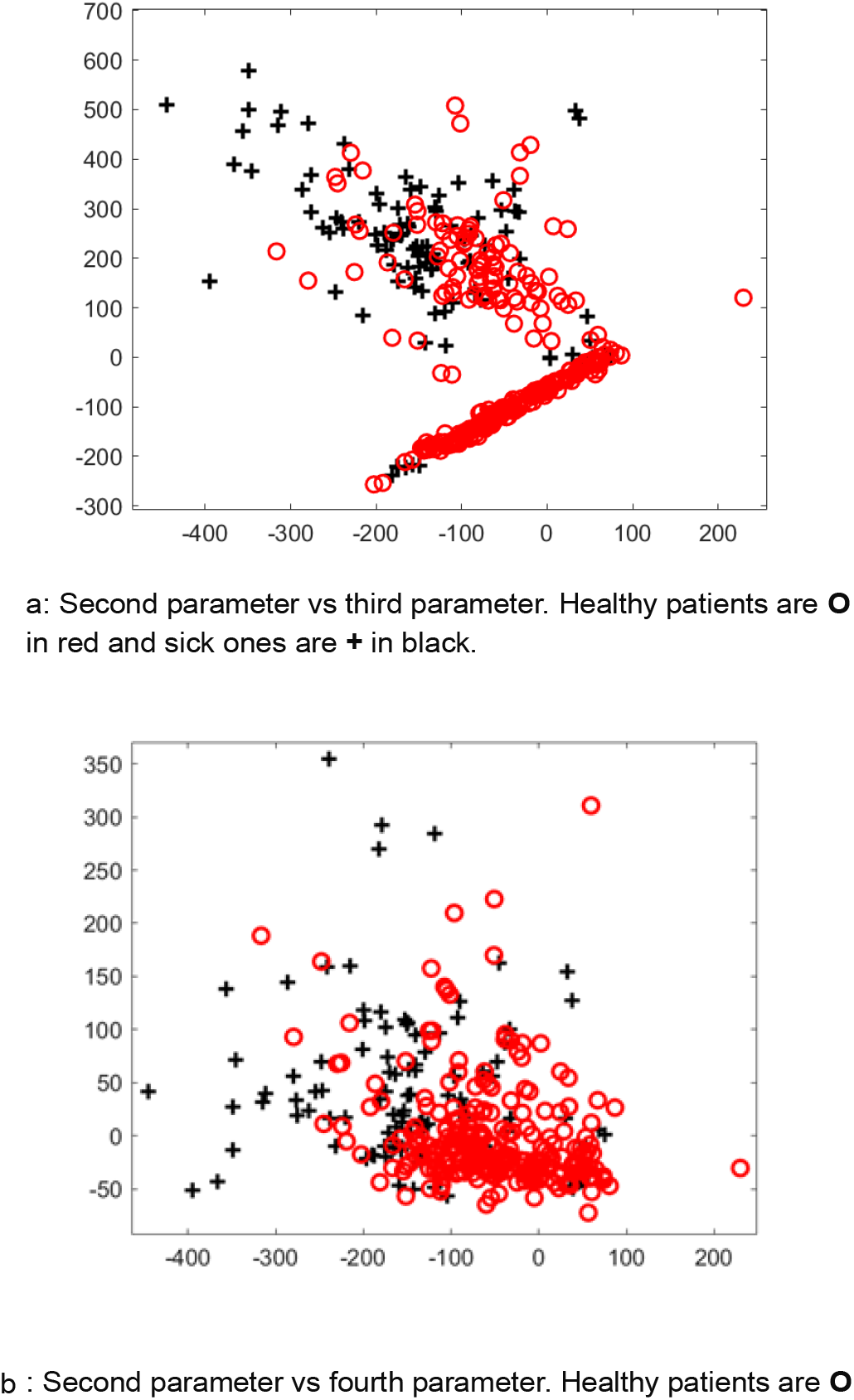

## IIII Discussion

### A. Image Analysis

Using DTI breast data, including a static frame, and applying PCA, we have separated the data into three independent images and three time traces. We identified the first image as the vascular system, the second as possibly related to venous vasodilation, and the third as a slow hormonal vascular response.

We have four patients with an identified tumor location. In two cases, we can detect tumor heat; in the other two, we can locate the vascular system blocking heat propagation at the expected tumor location. It appears that a hot image in a late response indicates the cancer location. It also seems that the dense vascular system between the tumor and the skin will mask it. More data is needed to support these conclusions. In the two cases blocked by the vascular system, it might be possible to perform reverse modelling using data away from the superficial vascular heat to identify a potential hidden source or to use AI. More data with identified cancer locations are needed to verify our conclusions.

### B Time Signature Analysis

The ICA algorithm produced two matrices, one for images and one for the three time traces. Understanding it requires additional analysis. Using ICA, we obtained four subcomponents. We used three to classify the patient as sick or healthy. At this stage, we can only speculate about the algorithm’s mechanism. The three traces contain information about the vascular system’s physiology. In a previous study [6], we identified and imaged vasomodulation of veins that depends on the presence of cancer [33][34][35]. Each trace in the static frame contains information about its static value. Applying VWT might reveal the mechanism behind the algorithm. Unlike AI, we can trace the process and figure it out.

A noteworthy fact is that we developed the algorithm across the entire data set without distinguishing between healthy and sick patients, because it figured out the separation on its own. The algorithm is designed to find anomalies, not cancer in particular. The algorithm identified one subvector common to both healthy and sick patients and three subvectors separating sick from healthy patients. Those three vectors were also present in part of the healthy population. When applying the algorithm to the entire population, as depicted in Figures 6 and 7, we observe a clear separation between the two subgroups.

The majority of healthy patients are in a straight-line group, and sick patients are distributed in a cloud-like pattern. We see some leakage between the groups. Six sick patients appear in the linear healthy group, and a larger number of healthy patients are located in the cloud, but in a subgroup closer to the linear healthy group. This effect was somewhat surprising, as we expected some mixing, but in the opposite direction. The original dataset’s classification of sick patients didn’t distinguish between the two breasts, so we expected half of them to be healthy. In this study, only six, with possibly three more, are in the healthy group. And four occurred just outside the linear group. A surprisingly large number of healthy patients appear in the sick cloud. In routine screening populations, about 0.5% of patients are diagnosed with cancer.

The results here are much higher. Those healthy patients form a subgroup located between the healthy linear group and the sick subgroup. This result isn’t random; the algorithm identified something we weren’t aware of — possibly a pre-cancerous stage that might or might not develop into active cancer. It is not clear what the algorithm flags out as an exception. It may detect a lack of vasodilation or excessive vascular heat. We should apply the tools developed in previous work [6] to identify those possibilities.

## IIIII Conclusions

Recent studies have focused on using AI with static thermal images to detect breast cancer, often reporting unrealistic accuracy. Our research takes a different, non-AI approach using dynamic thermal images and ICA. Our previous physics-based study using virtual waves successfully identified blood vessels and vaso dilation, but failed to detect cancer because the virtual waves did not register the static heat from tumors. The current work solves this problem by skipping the virtual wave step. Instead, we developed a two-layer algorithm (similar to AI) to identify the absence of vasodilation, a potential sign of cancer. Unlike AI’s “black box” algorithms, our method uses ICA as a “glass box,” producing highly interpretable images. ICA is reversible to a high degree. This reversibility allows us to understand the algorithm’s mechanism, which we plan to explore further in future work to create even more interpretable results.

A lack of evidence and interpretable data has created a 40-year barrier to the clinical adoption of an FDA-approved imaging modality by breast radiologists. The author presents a new method for actual patient data, providing visual correspondence. In earlier work[6], we identified the physiological response of the vascular system to external cooling. The initial goal of this work was to isolate blood vessels as a preprocessing step to identify deeper, static heat sources by thermal using ICA matrix factorization. We achieved this goal, but with some limitations due to the superficial vascular system occasionally masking the tumor heat.

During this work, we identified an algorithm based on the time trace of the matrix factorization H that separates patients into two subgroups: one with the majority of healthy patients and the other with the majority of sick patients. The algorithm classified some healthy patients as sick. Although the screening mechanism was not identified, this method is superior to AI, as it allows us to trace the algorithm and understand its decision tree. While this work was based on low-quality data, it requires validation with more reliable data. Better data, with sufficient characterization of the cancer, are necessary, particularly for healthy patients who fall outside the healthy linear subgroup.

## Data Availability

Raw data available
http://visual.ic.uff.br/dmi

## Acknowledgment

Thanks to Steven Fry for going over the manuscript.

